# Awareness-driven Behavior Changes Can Shift the Shape of Epidemics Away from Peaks and Towards Plateaus, Shoulders, and Oscillations

**DOI:** 10.1101/2020.05.03.20089524

**Authors:** Joshua S. Weitz, Sang Woo Park, Ceyhun Eksin, Jonathan Dushoff

## Abstract

The COVID-19 pandemic has caused more than 1,000,000 reported deaths globally, of which more than 200,000 have been reported in the United States as of October 1, 2020. Public health interventions have had significant impacts in reducing transmission and in averting even more deaths. Nonetheless, in many jurisdictions the decline of cases and fatalities after apparent epidemic peaks has not been rapid. Instead, the asymmetric decline in cases appears, in most cases, to be consistent with plateau- or shoulder-like phenomena – a qualitative observation reinforced by a symmetry analysis of US state-level fatality data. Here we explore a model of fatality-driven awareness in which individual protective measures increase with death rates. In this model, fast increases to the peak are often followed by plateaus, shoulders, and lag-driven oscillations. The asymmetric shape of model-predicted incidence and fatality curves are consistent with observations from many jurisdictions. Yet, in contrast to model predictions, we find that population-level mobility metrics usually increased from low early-outbreak levels *before* peak levels of fatalities. We show that incorporating fatigue and long-term behavior change can reconcile the apparent premature relaxation of mobility reductions and help understand when post-peak dynamics are likely to lead to a resurgence of cases.

## I INTRODUCTION

The spread of COVID-19 has elevated the importance of epidemiological models as a means to forecast both near- and long-term spread. In the United States, the Institute for Health Metrics and Evaluation (IHME) model has emerged as a key influencer of state- and national-level policy [1]. The IHME model includes a detailed characterization of the variation in hospital bed capacity, ICU beds, and ventilators between and within states. Predicting the projected strains on underlying health resources is critical to supporting planning efforts. However such projections require an epidemic ‘forecast’. Early versions of IHME’s epidemic forecast differed from conventional epidemic models in a significant way – IHME assumed that the cumulative deaths in the COVID-19 epidemic followed a symmetric, Gaussian-like trajectory. For example, the IHME model predicted that if the peak is 2 weeks away then in 4 weeks cases will return to the level of the present, and continue to diminish rapidly. But, epidemics need not have one symmetric peak – the archaic Farr’s Law of Epidemics notwithstanding (see [2] for a cautionary tale of using Farr’s law as applied to the HIV epidemic).

Conventional epidemic models of COVID-19 represent populations in terms of their ‘status’ vis a vis the infectious agent, i.e., susceptible, exposed, infectious, hospitalized, and recovered [3–9]. New transmission can lead to an exponential increases in cases when the basic reproduction number ℛ_0_ *>* 1 (the basic reproduction number denotes the average number of new infections caused by a single, typical individual in an otherwise susceptible population [10]). Subsequent spread, if left unchecked, would yield a single peak – in theory. That peak corresponds to when ‘herd immunity’ is reached, such that the effective reproduction number, ℛ_eff_ = 1. The effective reproduction number denotes the number of new infectious cases caused by a single infectious individual in a population with pre-existing circulation. But, even when herd immunity is reached, there will still be new cases which then diminish over time, until the epidemic concludes. A single-peak paradigm is robust insofar as the disease has spread sufficiently in a population to reach and exceed ‘herd immunity’. The converse is also true – as long as a population remains predominantly immunologically naive, then the risk of further infection has not passed.

In contrast to the IHME model, the Imperial College of London (ICL) model [3] used a conventional state-driven epidemic model to show the benefits of early intervention steps in reducing transmission and preserving health system resources vs. a ‘herd immunity’ strategy. The ICL model assumed that transmission is reduced because of externalities, like lockdowns, school closings, and so on. As a result, early predictions of the ICL model suggested that lifting of large-scale public health interventions could be followed by a second wave of cases. This has turned out to be the case, in some jurisdictions. Yet, for a disease that is already the documented cause of more than 200,000 deaths in the United States alone, we posit that individuals are likely to continue to modify their behavior even after lockdowns are lifted. Indeed, the peak death rates in the United States and globally are not as high as potential maximums in the event that COVID-19 had spread unhindered in the population [3]. Moreover, rather than a peak and symmetric decline, there is evidence of asymmetric plateaus and shoulder-like behavior for daily fatality rates in the spring-summer trajectory of the pandemic in US-states (Figure 1; full state-level data in SI Appendix, Fig. S1). These early plateaus have been followed, in many cases, with resurgence of cases and fatalities.

**FIG. 1:**
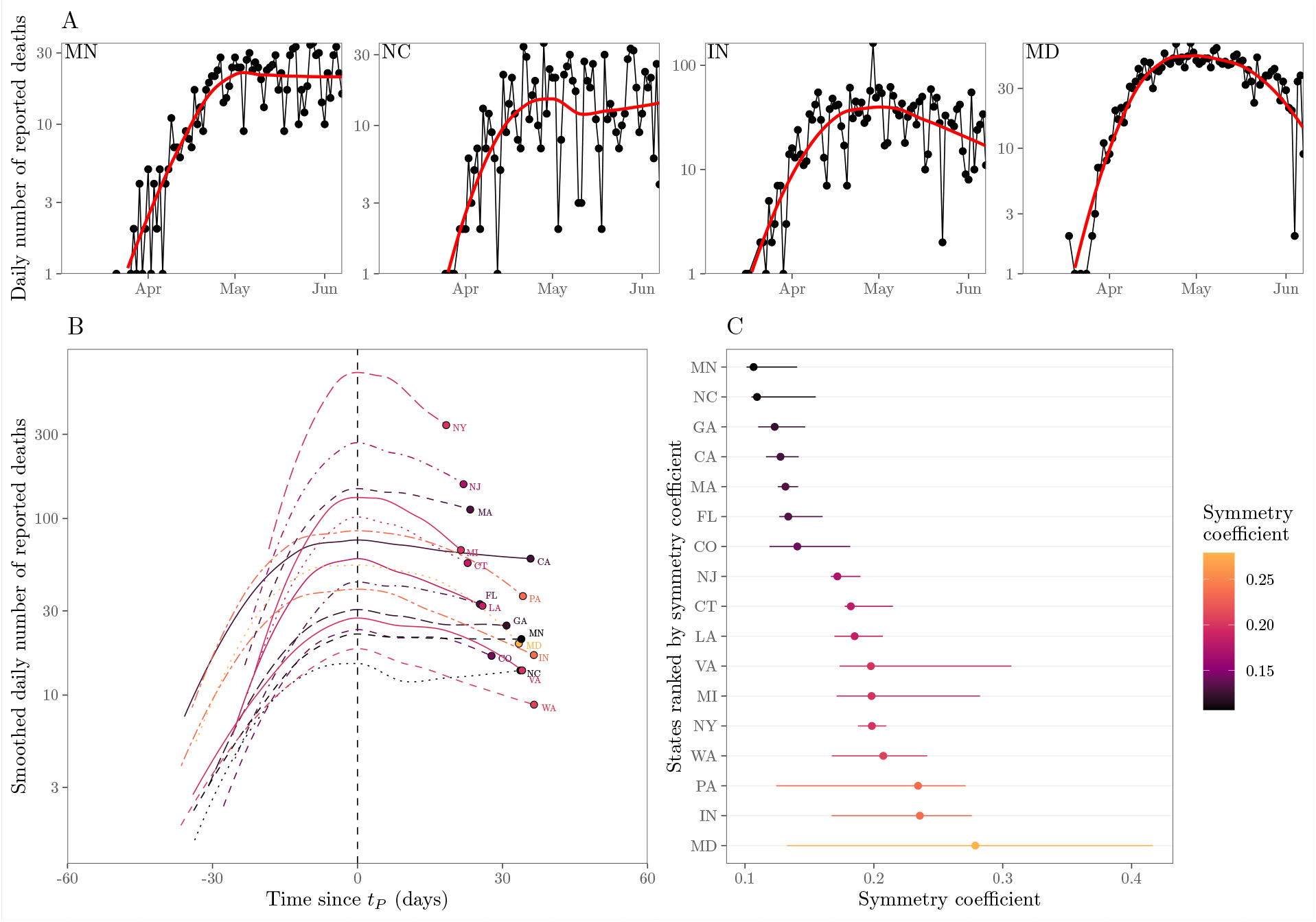
Plateaus and shoulder-like dynamics in COVID-19 fatalities. (A) Examples of daily number of reported deaths for COVID-19 (black points and lines) and the corresponding locally estimated scatterplot smoothing (LOESS) curves (red lines) in four states, including two estimated to be the most plateau-like (Minnesota and North Carolina) and two estimated to be the most peak-like (Indiana and Maryland). Daily number of deaths is smoothed in log space, only including days with one or more reported deaths. We restrict our analysis to states in which the peak smoothed death is greater than 10 as of June 7, 2020 (resulting in 17 states in total). (B) Smoothed daily number of reported deaths centered around the first peak time *t*_*P*_ across 17 states. Smoothed death curves are plotted between *t*_*P*_ − Δ*t* and *t*_*P*_ + Δ*t*, where Δ*t* is defined such that smoothed death at time *t*_*P*_ − Δ*t* corresponds to 10% of the smoothed peak value. (C) Measured symmetry coefficient and confidence intervals. Symmetry coefficient is calculated by dividing the death value at time *t*_*P*_ − Δ*t* by the death value at time *t*_*P*_ + Δ*t*. If the death curve is symmetric, the symmetry coefficient should equal 1. Confidence intervals are calculated by bootstrapping across the date of deaths for each individual 1000 times and recalculating the symmetry coefficient (after smoothing each bootstrap time series). LOESS smoothing is performed by using the `loess` function in R.

In this manuscript we use a nonlinear model of epidemiological dynamics to ask the question: what is the anticipated shape of an epidemic if individuals modify their behavior in direct response to the impact of a disease at the population level? In doing so, we build upon earlier work on awareness based models (e.g. [11–14]) with an initial assumption: individuals reduce interactions when death rates are high and increase interactions when death rates are low. As we show, short-term awareness can lead to dramatic reductions in death rates compared to models without accounting for behavior, leading to plateaus, shoulders, and lag-driven oscillations in death rates. We also show that dynamics can be driven from persistent dynamics to elimination when awareness shifts from short- to long-term. Notably, we find that despite model predictions, the empirical data reveals that mobility increased even as fatalities were increasing. This reveals the potential role for fatigue and long-term changes in behavior beyond those linked to mobility (e.g., mask-wearing) in shaping Covid-19 dynamics.

## II RESULTS AND DISCUSSION

### A SEIR Model with Short-Term Awareness of Risk

Consider an SEIR like model

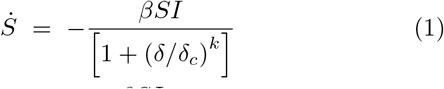

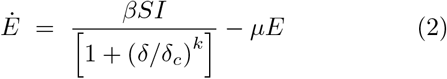

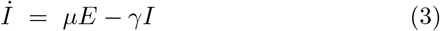

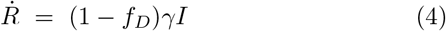

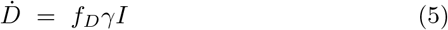

where *S, E, I, R*, and *D* denote the proportions of susceptible, exposed, infectious, recovered, and deaths, respectively, given transmission rate *β* /day, transition to infectious rate *µ* /day, recovery rate *γ* /day, where *f*_*D*_ is the infection fatality probability. The awareness-based distancing is controlled by the death rate 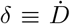, the half-saturation constant (*δ*_*c*_ *>* 0), and the sharpness of change in the force of infection (*k* ≥1) (see Figure 2 for a schematic and Table I for a list of all parameters used in models). Since *δ* is proportional to *I*, this model is closely related to a recently proposed awareness-based distancing model [14] and to an independently derived feedback SIR model [15]. Note that the present model converges to the conventional SEIR model as *δ*_*c*_ → ∞.

**TABLE I:** Parameter descriptions and values/ranges used for simulations. Transmission rate is chosen to match *ℛ*_0_ = 3.

| Notation | Description | Values/Ranges |
| --- | --- | --- |
| $\beta$ | Transmission rate | $1/2 \text{ days}^{-1}$ |
| $1/\mu$ | Mean latent period | 2 days |
| $1/\gamma$ | Mean infectious period | 6 days |
| $1/\gamma_H$ | Mean time in a hospital stay before a fatality | 7–28 days |
| $f_D$ | infection fatality probability | 0.01 |
| $N$ | Population size | $10^7$ |
| $N\delta_c$ | Half-saturation constant for short-term awareness | 5–500 deaths/day |
| $ND_c$ | Half-saturation constant for long-term awareness | 2,500–10,000 deaths |
| $k$ | sharpness of change in the force of infection | 1–4 |
| $\epsilon$ | Time scale of behavior change | $1/7 \text{ days}^{-1}$ |

**FIG. 2:**
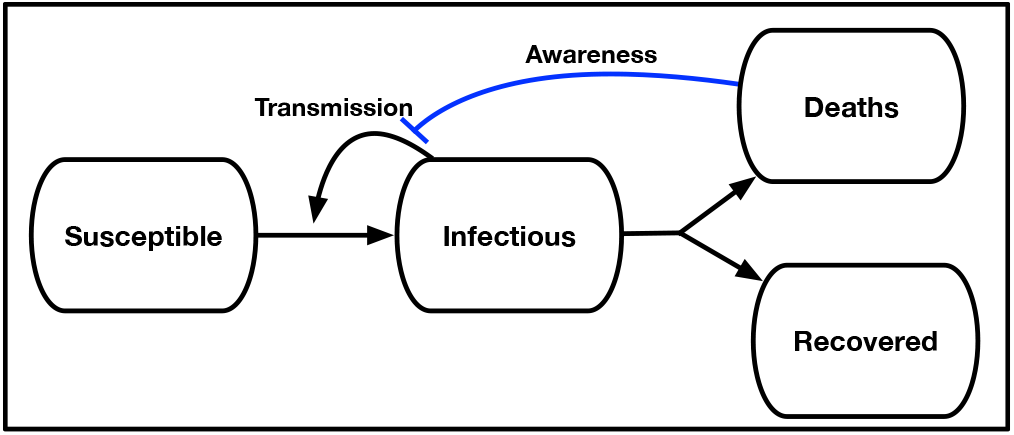
Schematic of an SEIR model with awareness-driven social distancing. Transmission is reduced based on short- and/or long-term awareness of population-level disease severity (i.e., fatalities).

Uncontrolled epidemics in SEIR models have a single case peak, corresponding to the point where *γI* = *βSI* such that the population obtains herd immunity when only a proportion *S* = 1*/*ℛ_0_ have yet to be infected. However, in the model above individuals decrease transmission in response to awareness of the impacts of the disease, *δ*(*t*). In this case, infected cases can peak even when the population is far from herd immunity, specifically when

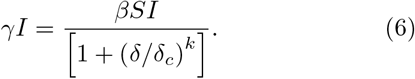

When *δ*_*c*_ is small compared to the per-capita death rate of infectious individuals (*γf*_*D*_) we anticipate that individual behavior will respond quickly to the disease outbreak. Hence, we hypothesize that the emergence of an awareness-based peak can occur early, i.e., *S*(*t*) ≈1, consistent with a quasi-stationary equilibrium when the death rate is

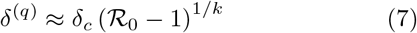

and the infection rate is

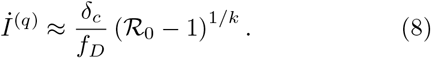

These quasi-equilibrium is maintained not because of herd immunity, but because of changes in behavior.

We evaluate this hypothesis in Figure 3 for *k* = 1, *k* = 2, and *k* = 4 given disease dynamics with *β* = 0.5/day, *µ* = 1*/*2/day, *γ* = 1*/*6/day, *f*_*D*_ = 0.01, *N* = 10^7^, and *Nδ*_*c*_ = 50/day. As is evident, the rise and decline from peaks are not symmetric. Instead, incorporating awareness leads to dynamics where incidence decreases very slowly after a peak. The peaks occur at levels of infection far from that associated with herd immunity. Post-peak, shoulders and plateaus emerge because of the balance between relaxation of awareness-based distancing (which leads to increases in cases and deaths) and an increase in awareness in response to increases in cases and deaths. As the steepness of response *k* increases, individuals become less sensitive to fatality rates where *δ < δ*_*c*_ and more sensitive to fatality rates where *δ > δ*_*c*_. This leads to sharper dynamics. In addition, infections can over-shoot the expected plateau given that awareness is driven by fatalities which are offset with respect to new infections.

**FIG. 3:**
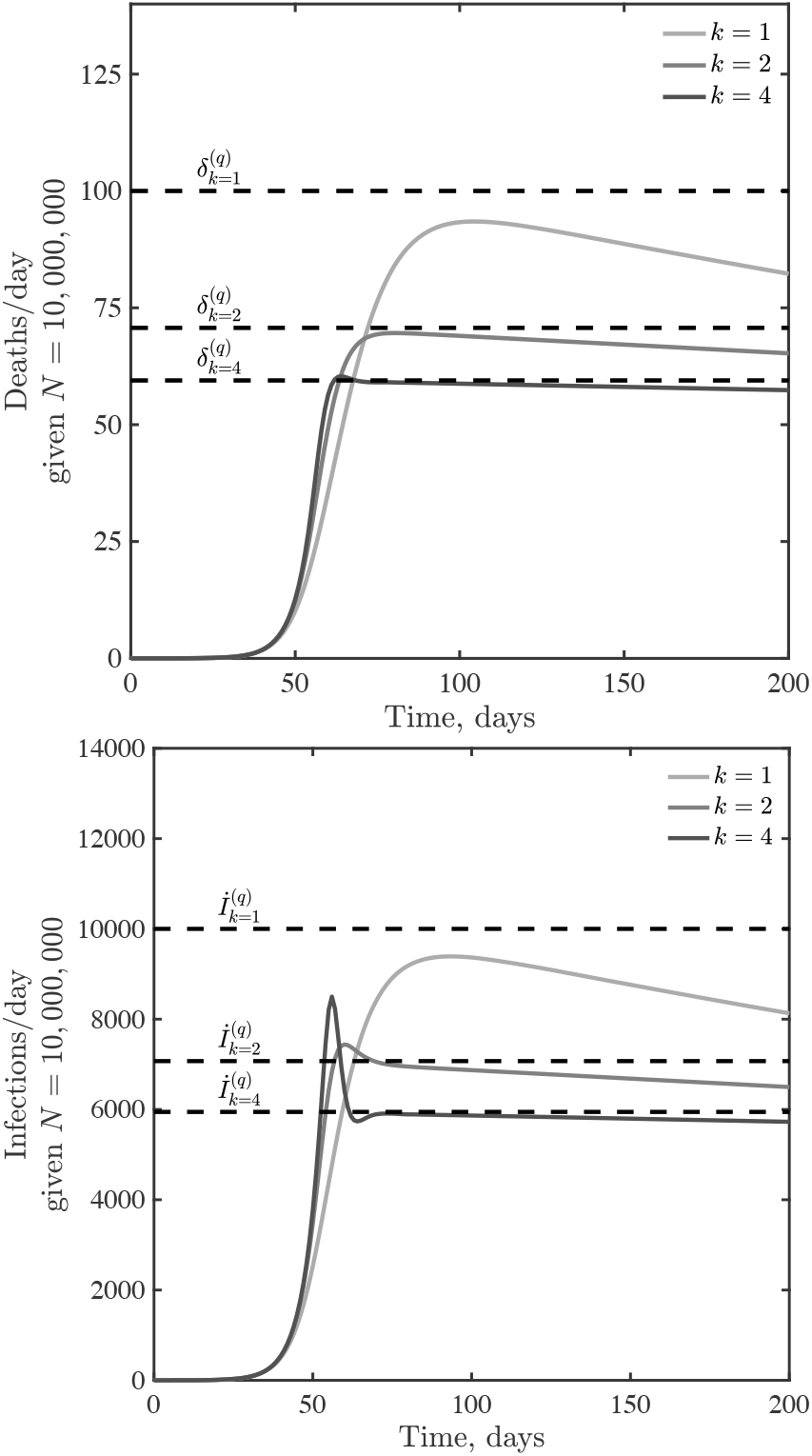
Infections and deaths per day in a death-awareness based social distancing model. Simulations have the epidemiological parameters *β* = 0.5 /day, *µ* = 1*/*2 /day, *γ* = 1*/*6/day, and *f*_*D*_ = 0.01, with variation in *k* = 1, 2 and 4. We assume *Nδ*_*c*_ = 50 /day in all cases.

## B Short-term awareness, long-term plateaus, and oscillations

Initial analysis of an SEIR model with short-term awareness of population-level severity suggests a generic outcome: fatalities will first increase exponentially before before slowing to plateau at a level near *δ*_*c*_. Figure 4 shows dynamics for values of *δ*_*c*_ ranging from to 5 to 500 deaths/day in a population of 10^7^ (here *k* = 2; results for *k* = 1 or *k* = 4 are similar, see SI Appendix, SI Fig. S2). When *δ*_*c*_ is small (compared to (*γf*_*D*_), fatalities can be sustained at near-constant levels for a long time. When *δ*_*c*_ is higher then the decline of cases and fatalities due to susceptible depletion is relatively fast. However, over a wide range of assumptions about critical daily fatality rates *δ*_*c*_, the population remains largely susceptible even as sustained fatalities continue for a period far greater than the time it took to reach the plateau. To explore the impacts of lags on dynamics, we incorporated an additional class *H*, assuming that fatalities follow potentially prolonged hospital stays. We do not include detailed information on symptomatic transmission, asymptomatic transmission, hospitalization outcome, age structure, and age-dependent risk (as in [3]). Instead, we consider the extended SEIR model:

**FIG. 4:**
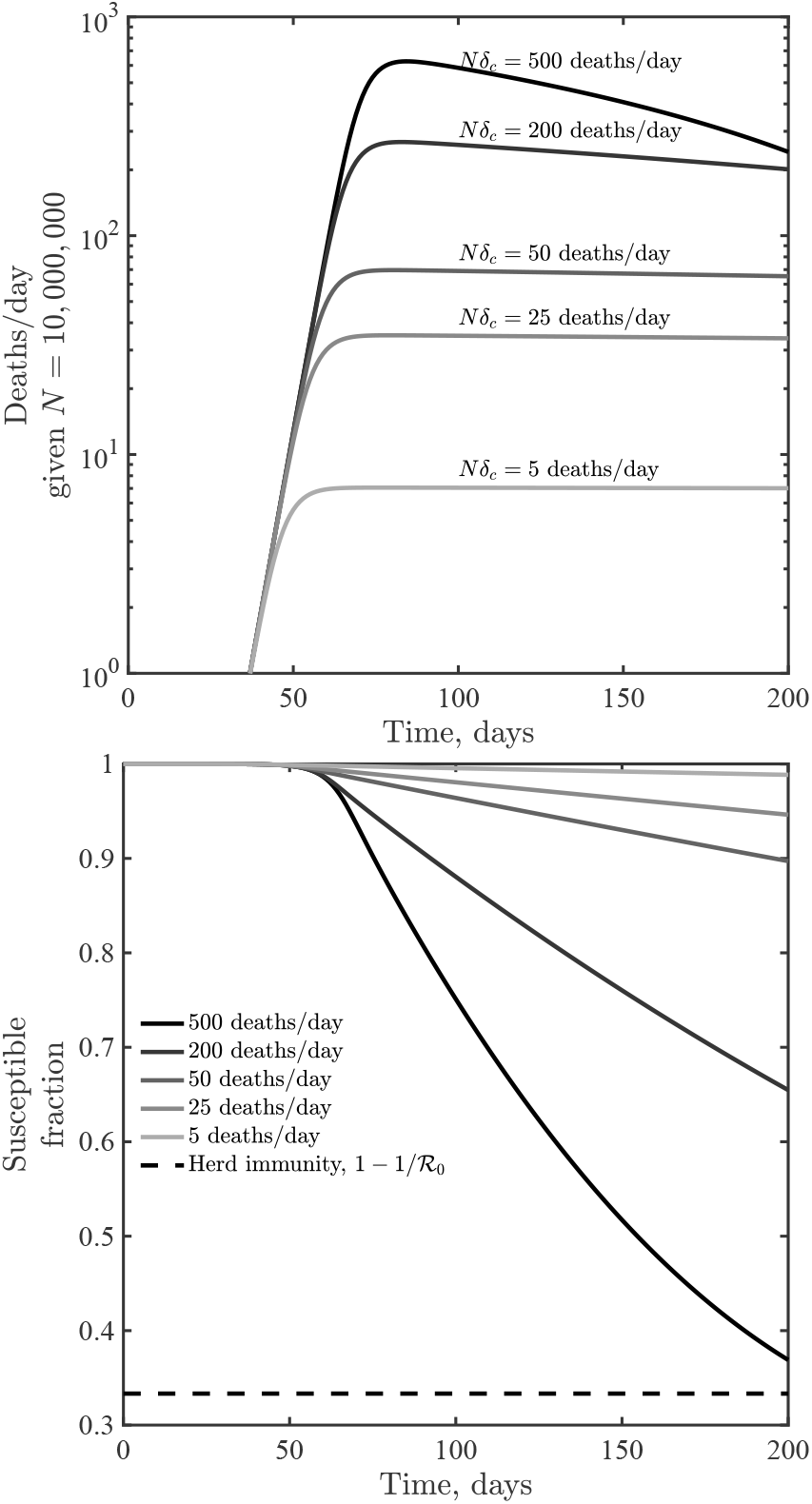
Dynamics given variation in the critical fatality awareness level, *δ*_*c*_ for awareness *k* = 2. Panels show deaths/day (top) and the susceptible fraction as a function of time (bottom), the latter compared to a herd immunity level when only a fraction 1*/ R* 0 remain susceptible. These simulations share the epidemiological parameters *β* = 0.5 /day, *µ* = 1*/*2 /day, *γ* = 1*/*6 /day, and *f*_*D*_ = 0.01.

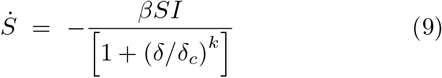

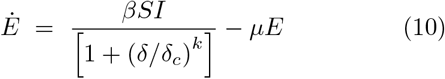

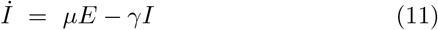

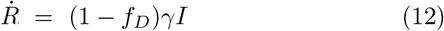

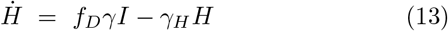

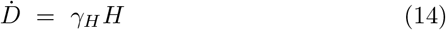

where *T*_*H*_ = 1*/γ*_*H*_ defines the average time in a hospital stay before a fatality. Note, we recognize that many individuals recover from COVID-19 after hospitalization; this model’s hospital compartment functions as a prefilter.

The earlier analysis of the quasi-stationary equilibrium in fatalities holds in the case of a SEIR model with additional classes before fatalities. Hence, we anticipate that dynamics should converge to *δ* = *δ*^(*q*)^ at early times. However, increased delays between cases and fatalities could lead to oscillations. Indeed, this is what we find via examination of models in which *T*_*H*_ ranges from 7 to 28 days, with increasing magnitude of oscillations as *T*_*H*_ increases (see Figure 5 for *k* = 2 with qualitatively similar results for *k* = 1 and *k* = 4 shown in SI Appendix, SI Fig. S3).

**FIG. 5:**
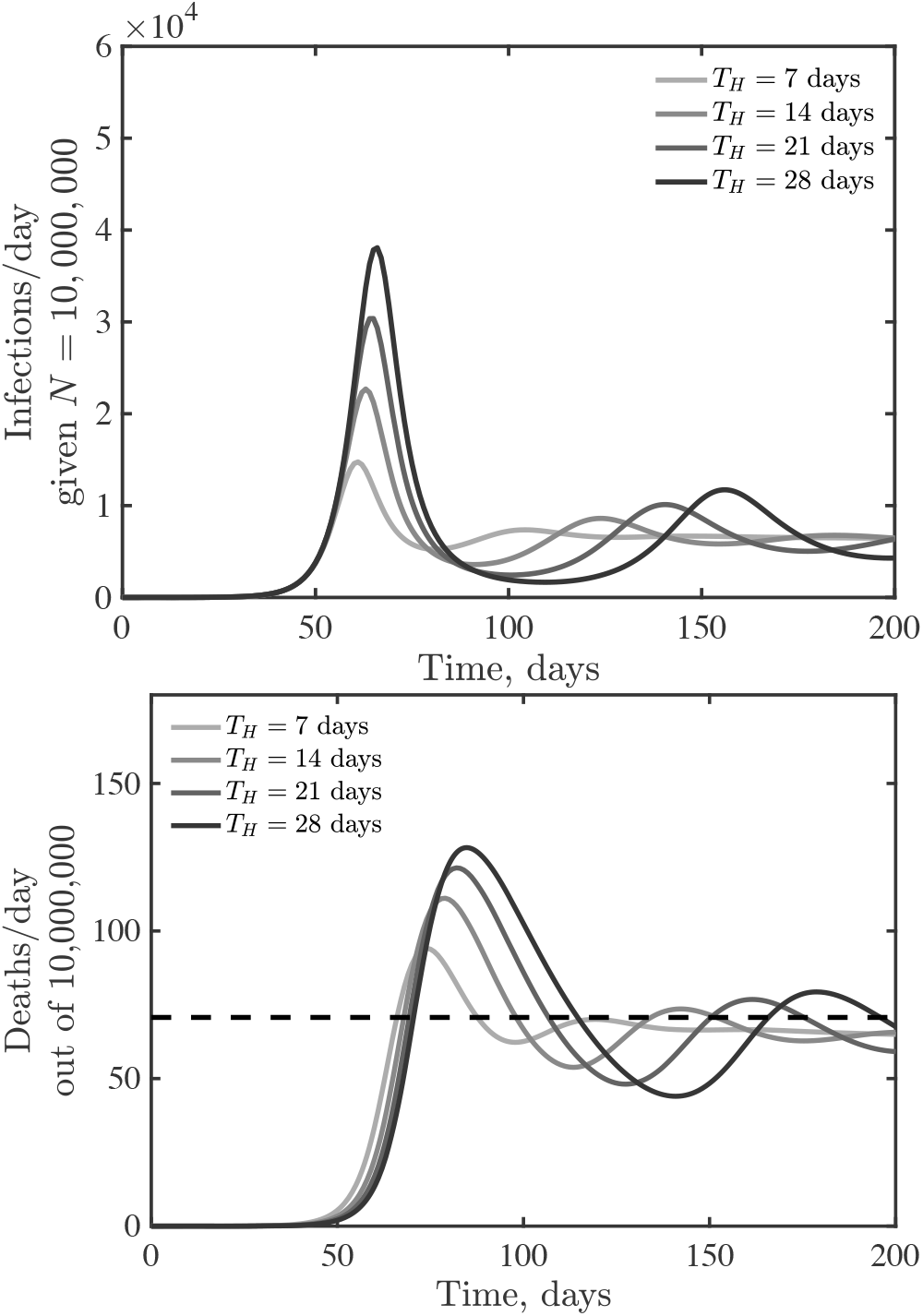
Emergence of oscillatory dynamics in a death-driven awareness model of social distancing given lags between infection and fatality. Awareness is *k* = 2 and all other parameters as in Figure 3. The dashed lines for fatalities expected quasi-stationary value *δ*^(*q*)^.

## C Dynamical consequences of short-term and long-term awareness

Awareness can vary in duration, e.g., awareness of SARS-CoV-2 may prepare individuals to more readily adopt and retain social distancing measures [16, 17]. In previous work, long-term awareness of cumulative incidence was shown to lead to substantial decreases in final size of epidemics compared to baseline expectations from inferred strength [14]. Hence, we consider an extension of the SEIR model with lags between infection and fatalities that incorporates both short-term and long-term awareness:

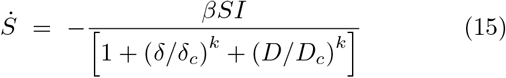

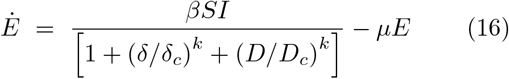

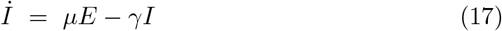

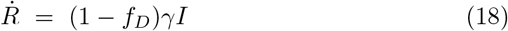

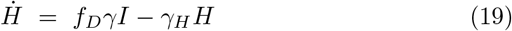

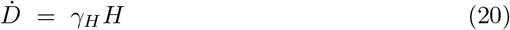

where *D*_*c*_ denotes a critical cumulative fatality level (and formally a half-saturation constant for the impact of longterm awareness on distancing). Note that the relative importance of short- and long-term awareness can be modulated by *δ*_*c*_ and *D*_*c*_ respectively. Figure 6 shows daily fatalities (top) and cumulative fatalities (bottom) for an SEIR model with ℛ_0_ = 2.5, *T*_*H*_ = 14 days, and *Nδ*_*c*_ = 50 fatalities per day and critical cumulative fatalities of *ND*_*c*_ = 2, 500, 5,000, 10,000 as well as a comparison case with vanishing long-term awareness. As is evident, long-term awareness drives dynamics towards rapid declines after reaching a peak. This decline arises because *D* monotonically increases; increasing fatalities beyond *D*_*c*_ leads to rapid suppression of transmission. However, when short-term, rather than long-term, awareness drives dynamics, then shoulders and plateaus can re-emerge. In reality, we expect that individual behavior is shaped by short- and long-term awareness of risks, including the potential for fatigue and ‘decay’ of long-term behavior change [11, 12].

**FIG. 6:**
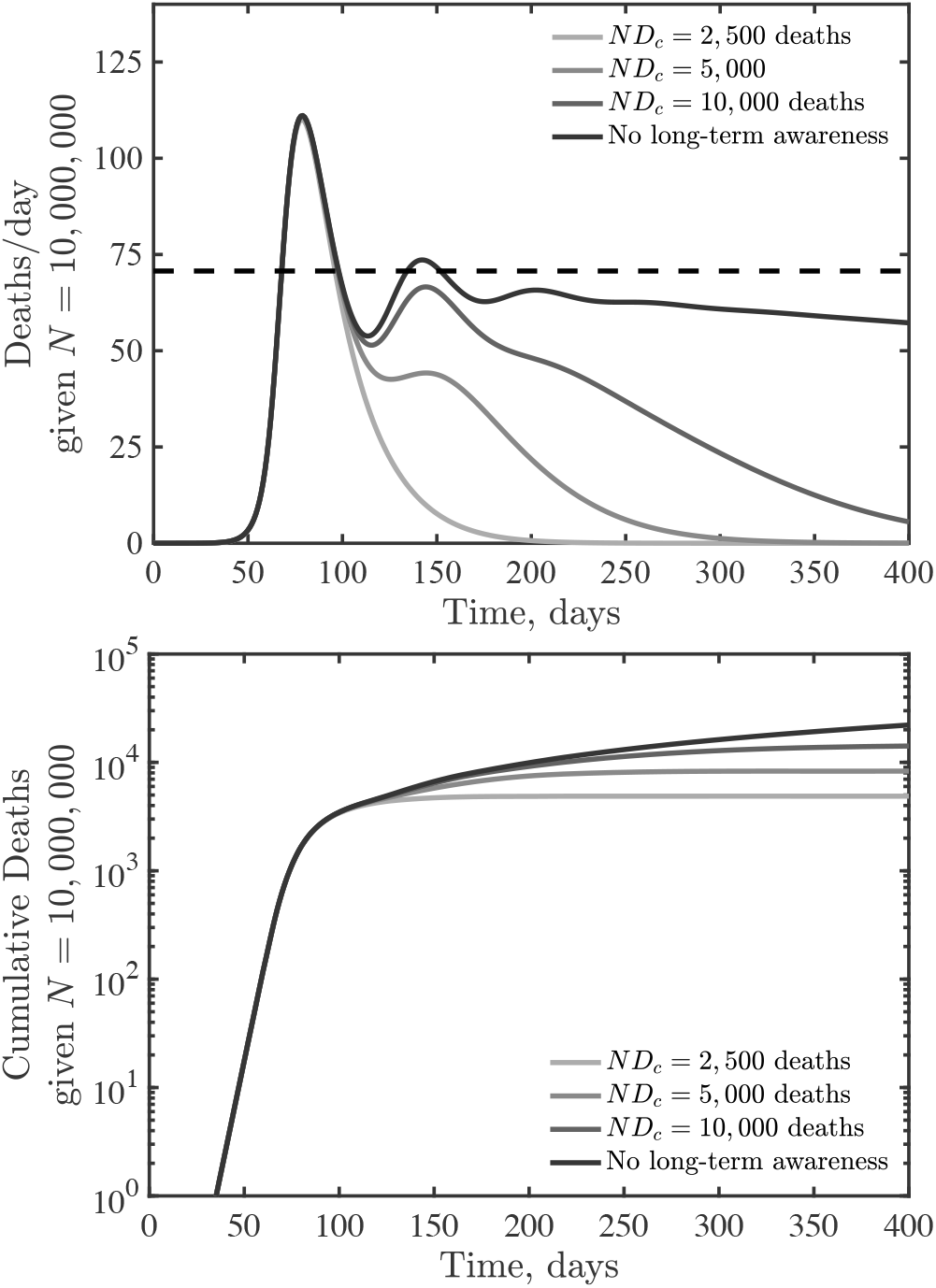
SEIR dynamics with short- and long-term awareness. Model parameters are *β* = 0.5 /day, *µ* = 1*/*2 /day, *γ* = 1*/*6 /day, *T*_*H*_ = 14 days, *f*_*D*_ = 0.01, *N* = 10^7^, *k* = 2, *Nδ*_*c*_ = 50 /day (short-term awareness), with varying *ND*_*c*_ (long-term awareness) as shown in the legend. The dashed line (top) denotes *δ*^(*q*)^ due to short-term distancing alone.

## D Empirical assessment of mechanistic drivers of asymmetric peaks in Covid-19 death rates

The models developed here suggest that awareness-driven distancing can lead to asymmetric epidemic peaks even in the absence of susceptible depletion. To test this hypothesis mechanistically, we jointly analyzed the dynamics of fatality rates and behavior, using mobility data obtained from Google COVID-19 Community Mobility Reports (https://www.google.com/covid19/mobility/) as a proxy for behavior (see Methods for the aggregation of multiple mobility metrics via a Principal Component Analysis (PCA)). Notably, we find that in the bulk of states examined aggregated rates of mobility typically began to *increase* before the local peak in fatality was reached (Figure 7A). This rebound in mobility rates implies that real populations are opening up faster than our simple model could predict. Awareness-driven models, shown in Figure 7B, show either ‘reversible’ or ‘counter-clockwise’ dynamics. In these models, risky behavior decreases until fatalities reach their peak. Models with short-term awareness but no long-term awareness exhibit a tight link between fatality and behavior (reversible behavior, like the top curve in Figure 7B). Models with long-term awareness exhibit counter-clockwise dynamics as risky behavior remains at low levels even as fatalities decrease; the asymmetry here is driven by the extent of long-term awareness.

**FIG. 7:**
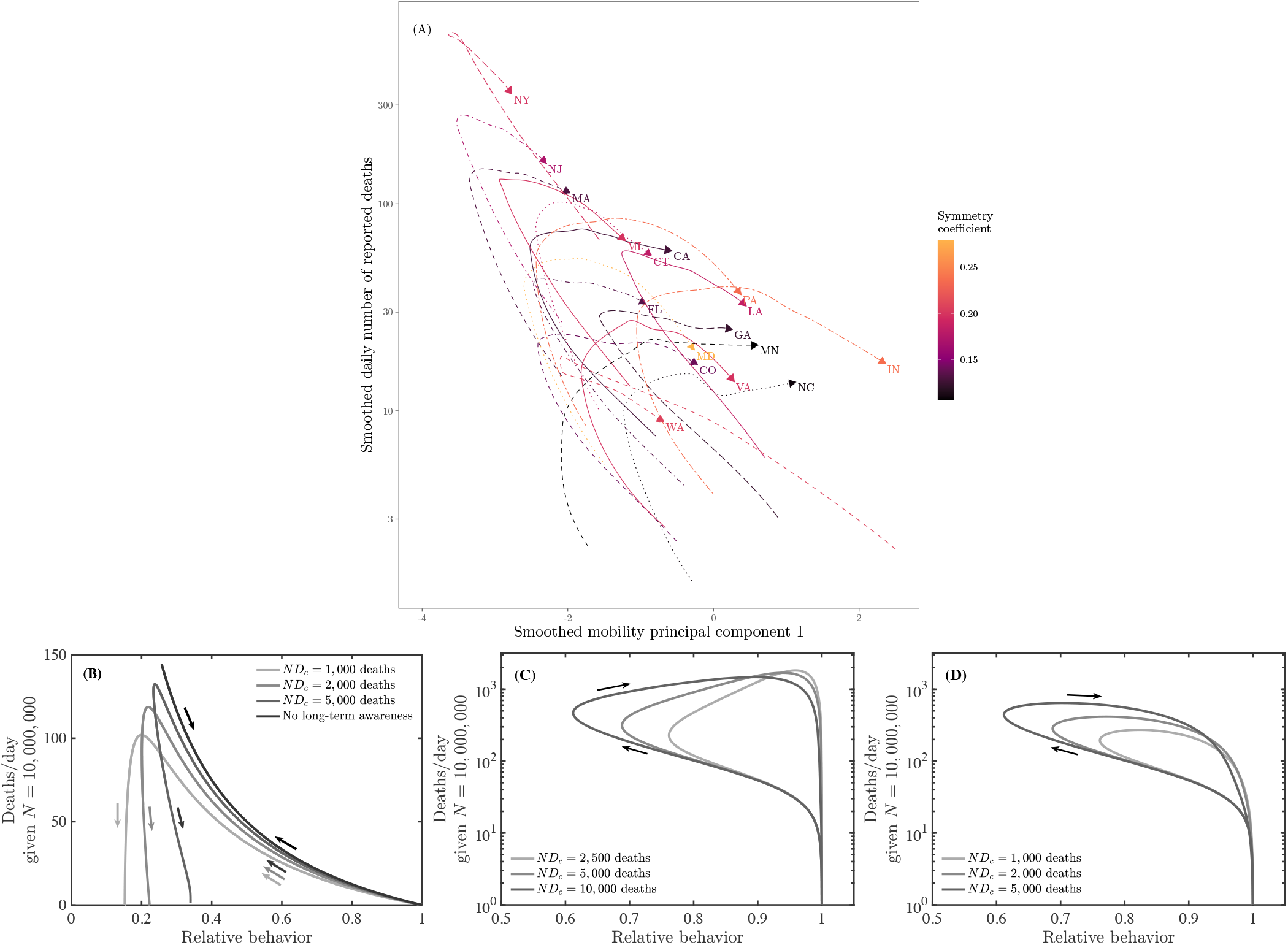
Phase-plane visualizations of deaths vs. mobility for state-level data (top) and SEIR models (bottom panels). (Top) Deaths and mobility indexes through time for the 17 analyzed states. Both data series are smoothed. Time windows as in 1. (B) Dynamics of effective behavior and death rates in a SEIR model with short- and long-term awareness. Curves denote different assumptions regarding long-term awareness, in each case *β* = 0.5/day, *µ* = 0.5/day, *γ* = 1*/*6/day, such that *R*0 = 3, with *k* = 2, *γ*_*H*_ = 1*/*21/day, and *f*_*D*_ = 0.01. The short-term awareness corresponds go *Nδ*_*c*_ = 50 deaths/day. Thin lines denote full dynamics over 400 days; thick lines denote the dynamics near the case fatality peak. (C) Dynamics of effective behavior and death rates in a SEIR model with awareness and fatigue. The three different curves denote different assumptions regarding long-term awareness, in each case *β* = 0.5/day, *µ* = 0.5/day, *γ* = 1*/*6/day, such that *R*0 = 3, with *k* = 2, *γ*_*H*_ = 1*/*21/day, *f*_*D*_ = 0.01, and *E* = 1*/*7/day. The short-term awareness corresponds go *Nδ*_*c*_ = 50 deaths/day. The force of infection does not include long term changes in behavior beyond mobility, i.e., *g*(*D*) = 1. (D) As in (C), but the force of infection includes long-term changes in behavior, i.e., *g*(*D*) = 1*/* (1 + (*D/D*_*c*_)^*k*^).

In contrast, the real data (Figure 7A) exhibit predominantly clockwise dynamics; exceptions include New York which has a nearly reversible (but still clockwise) pattern and Washington which has a counter-clockwise pattern anticipated by the awareness model. We hypothesized that a combination of awareness-driven distancing and fatigue could lead to clockwise dynamics: if people become fatigued with distancing behavior, then risk could rise even as deaths were rising. We developed the following model as a proof of concept in which fatigue is driven directly by deaths (though alternatives could also be explored linked to cases, hospitalizations, deaths and/or a combination):

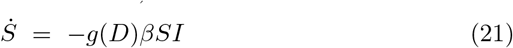

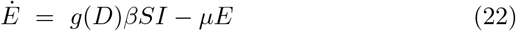

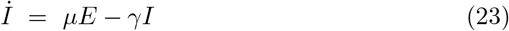

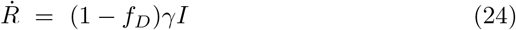

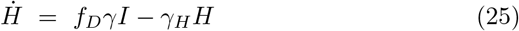

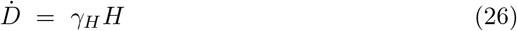

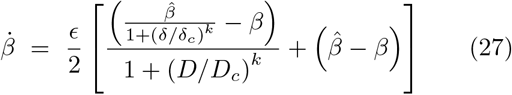

In this model with fatigue, the force of infection is related to the mobility denoted by *β*(*t*) (which dictates the number of interactions per unit time) modulated by a reduction in risk per infection *g*(*D*). In this model, 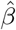 denotes the baseline behavior, and *ϵ* denotes a time-scale for behavior change. The level of fatigue is controlled by *D*_*c*_, such that mobility returns to a baseline 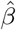 once *D* ≫ *D*_*c*_. We consider two models, corresponding to *g*(*D*) = 1 such that the force of infection depends on mobility alone, and *g*(*D*) = 1*/* (1 + (*D/D*_*c*_)^*k*^) corresponding to sustained changes in the risk of infection per contact (e.g., due to mask wearing, contact-less interactions, use of PPE, etc.). As shown in Figure 7C/D, the dynamics switch from counter-clockwise to clockwise in the *δ* − *β* plane given the incorporation of fatigue. Deaths drive down mobility, but eventually, decreases in *β* due to short-term awareness are over-come by fatigue, leading to increases in *β*. If *g*(*D*) = 1, then the dynamics include increases in both mobility and fatalities akin to levels expected in the absence of behavior, and eventually levels of infection that are stopped by herd immunity, rather than by awareness (see Figure 7C). In contrast, if there is sustained behavior change such that *g*(*D*) decreases with increasing cumulative deaths then there is a single peak that forms a clockwise loop; with the peak close to, but after the minimum in behavior (Figure 7D); as observed in nearly all state-level data sets.

## III CONCLUSIONS

We have developed and analyzed a series of models that assume awareness of disease-induced death can reduce transmission, and shown that such awareness-driven feedback can lead to highly asymmetric epidemic curves. Resulting fatality time series can exhibit extended periods of near-constant levels even as the majority of the population remains susceptible. Hence: passing a ‘peak’ need not imply the rapid decline of risk. In these conditions, if individuals are unable to sustain social distancing policies, or begin to tolerate higher death rates, then cases and fatalities could increase (similar results have also been proposed in a recent, independently derived feedback SIR model [15]). Indeed, detailed analysis of mobility and fatalities suggest that mobility increased before fatalities peaked with the exceptions of Washington and New York. This increase of mobility amidst rising fatalities is not consistent with simple models of awareness-driven distancing, but is consistent with more detailed models that include fatigue. Notably, we find that if mobility increases but the risk of infection per interaction decreases due to systemic changes in behavior, then models suggest ‘clockwise’ dynamics between behavior and fatality as found in nearly all state-level data sets analyzed here. Awareness-driven endogenous changes in ℛ_eff_ are typically absent in models that form the basis for public policy and strategic planning. Our findings highlight the potential impacts of short-term and long-term awareness in efforts to shape information campaigns to reduce transmission after early onset ‘peaks’, particularly when populations remain predominantly immunologically naive.

In moving from concept to intervention, it will be critical to address problems related to noise, biases, and the quantitative inference of mechanism from model-data fits. First, epidemic outbreaks include stochasticity of multiple kinds. Fluctuations could arise endogenously via process noise (especially at low levels of disease) or exogenously via time-varying parameters, Moreover, given the evidence for clustered transmission and super-spreading events [18–21], extensions of the present model framework should explicitly account for awareness-driven behavior associated with risky gatherings [22, 23]. Next, the link between severity and behavior change depends on reporting of disease outcomes. Biases may arise due to under-reporting of fatalities, particularly amidst intense outbreaks [24]. Awareness may also vary with community age structure and with other factors that influence the infection fatality rate and hence the link between total cases and fatalities [25]. Such biases could lead to systematic changes in awareness-driven responses. Finally, we recognize that estimating the influence of awareness-driven behavior change is non-trivial, given fundamental problems of identifiability. Nonetheless, it is important to consider the effects of behavior changes. Otherwise, reductions in cases (and fatalities) will necessarily be attributed to exogeneous factors (e.g., influential analyses of the impact of non-pharmaceutical interventions on Covid-19 in Europe do not account for changes in behavior [26]). Disentangling the impact of entangled interventions will require efforts to link model predictions with measurements of behavior, awareness, and disease dynamics.

Although the models here are intentionally simple, it seems likely that observed asymmetric dynamics of COVID-19, including slow declines and plateau-like behavior, is an emergent property of awareness-driven epidemiological dynamics. Moving forward, it is essential to fill in significant gaps in understanding how awareness of disease risk and severity shape behavior [27]. Mobility data is an imperfect proxy for distancing and other preventative behaviours, and thus for transmission risk. Thus far, measurements of community mobility have been used as a leading indicator for epidemic outcomes. Prior work has shown significant impacts of changes in mobility and behavior on the COVID-19 outbreak [7]. Here we have shown the importance of looking at a complementary feedback mechanism, i.e., from outbreak to behavior. In doing so, we have also shown that decomposing the force of infection in terms of the number of potential transmissions and the probability of infection per contact can lead to outcomes aligned with observed state-level dynamics. Understanding the drivers behind emergent plateaus observed at national and sub-national levels could help decision-makers structure intervention efforts appropriately to effectively communicate awareness campaigns that may aid in collective efforts to control the ongoing COVID-19 pandemic.

## IV METHODS

### A Epidemiological data

Daily number of reported deaths as of June 7, 2020, is obtained from The COVID Tracking Project (covid-tracking.com).

### B Mobility data

Mobility data as of June 12, 2020, are obtained from Google COVID-19 Community Mobility Reports (www.google.com/covid19/mobility/). The data set describes percent changes in mobility across six categories (grocery and pharmacy; parks; residential; retail and recreation; transit; and workplaces) compared to the median value from the 5–week period Jan 3–Feb 6, 2020. Raw mobility data are plotted in SI Appendix, SI Fig. S4.

### C Principal component analysis

We use principal component analysis (PCA) on the mobility data to obtain a univariate index of mobility. We exclude park visits from the analysis due to their anomalous, noisy patterns (SI Appendix, SI Fig. S4). Before performing PCA, we first calculate the 7-day rolling average for each mobility measure in order to remove the effects of weekly patterns. We combined mobility data from all 17 analyzed states, and standardized each measure (to zero mean and unit variance). The first principal component explains 93% of the total variance in this analysis, and the loading of the residential metric had a different sign from the other four mobility metrics. We thus used this component as our index of mobility (setting the direction so that only the residential metric contributed negatively to the index). To draw phase planes, we further smoothed our mobility index and daily reported deaths using locally estimated scatterplot (LOESS) smoothing. Daily number of deaths is smoothed in log space, only including days with one or more reported deaths. LOESS smoothing is performed by using the loess function in R.

## Data Availability

All simulation and codes used in the development of this manuscript are available at https://github.com/jsweitz/covid19-git-plateaus.

## Data availability

All simulation codes, figures, and data used in the development of this manuscript are available at https://github.com/jsweitz/covid19-git-plateaus.

## Acknowledgements

Research effort by JSW was enabled by support from grants from the Simons Foundation (SCOPE Award ID 329108), the Army Research Office (W911NF1910384), National Institutes of Health (1R01AI46592-01), and National Science Foundation (1806606 and 1829636). JD was supported in part by grants from the Canadian Institutes of Health Research and the Natural Sciences and Engineering Research Council of Canada.

## Appendix A Appendix - Supplementary Information

**FIG. S1:**
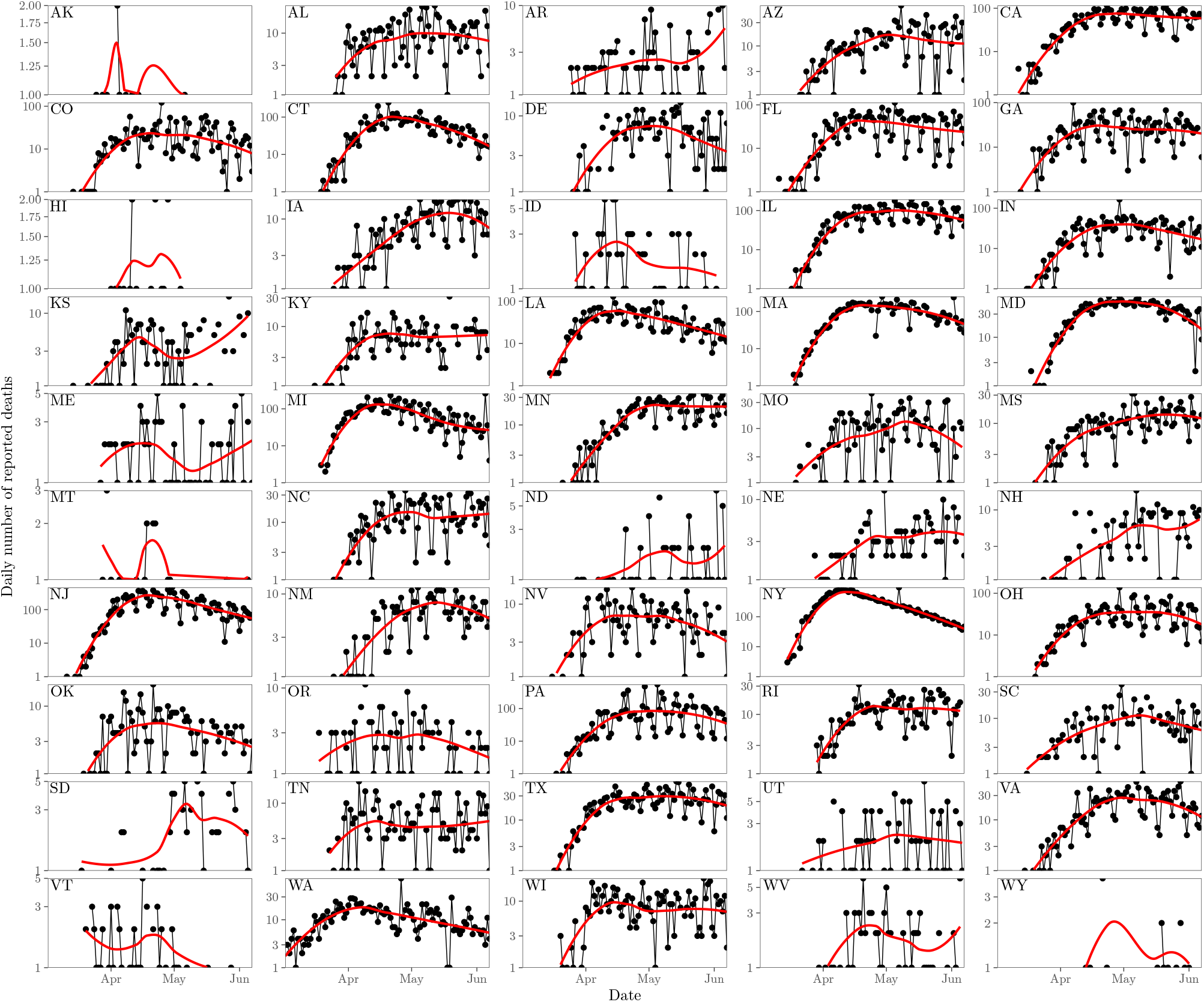
Daily number of reported deaths for COVID-19 (black points and lines) and the corresponding locally estimated scatterplot smoothing (LOESS) curves (red lines) in 50 states.

**FIG. S2:**
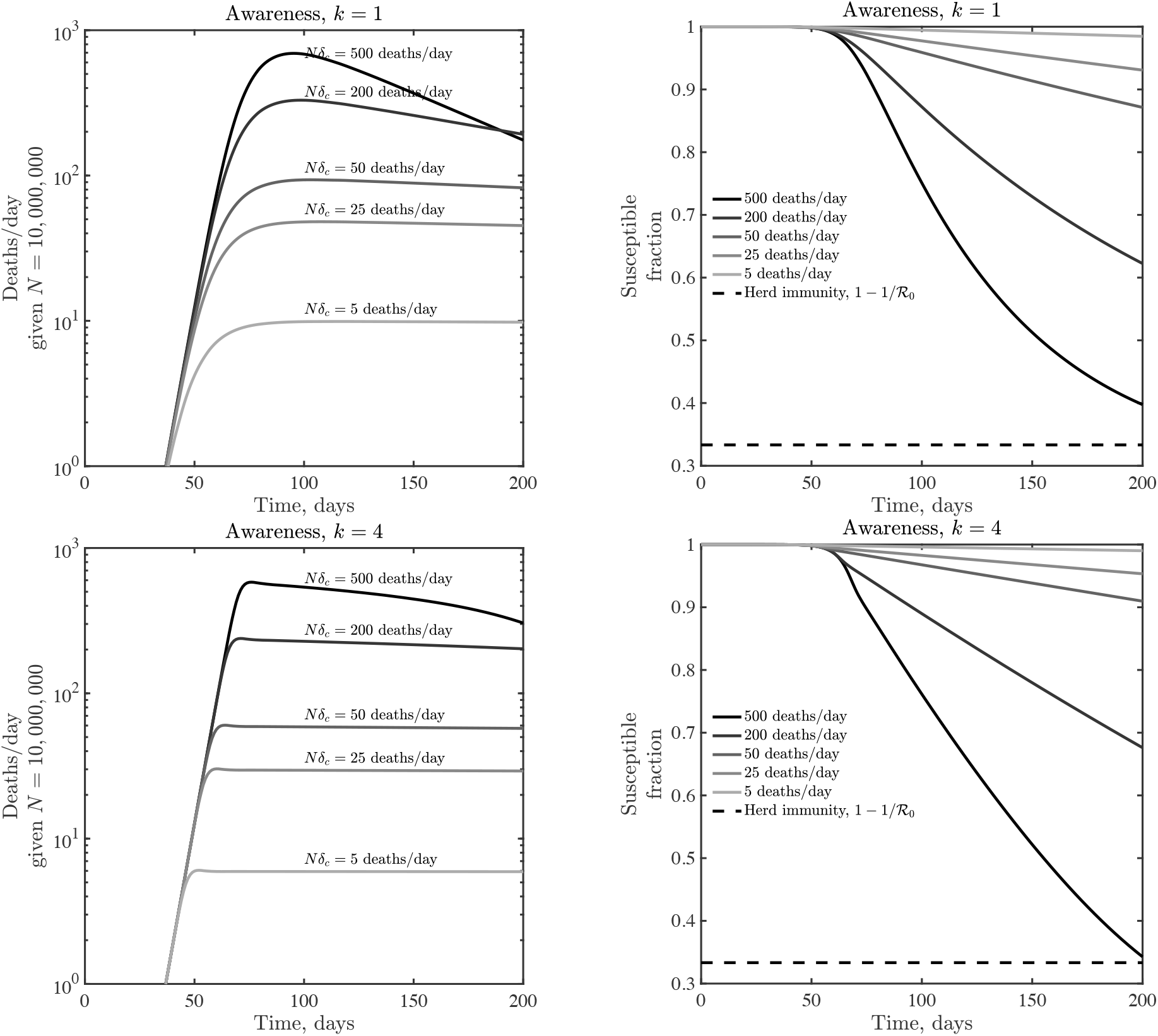
Dynamics given variation in the critical fatality awareness level, *δ*_*c*_ for awareness *k* = 1 (top) and *k* = 4 (bottom). Panels show deaths/day (top) and the susceptible fraction as a function of time (bottom), the latter compared to a herd immunity level when only a fraction 1*/ R*0 remain susecptible. These simulations share the epidemiological parameters *β* = 0.5 /day, *µ* = 1*/*2 /day, *γ* = 1*/*6 /day, and *f*_*D*_ = 0.01.

**FIG. S3:**
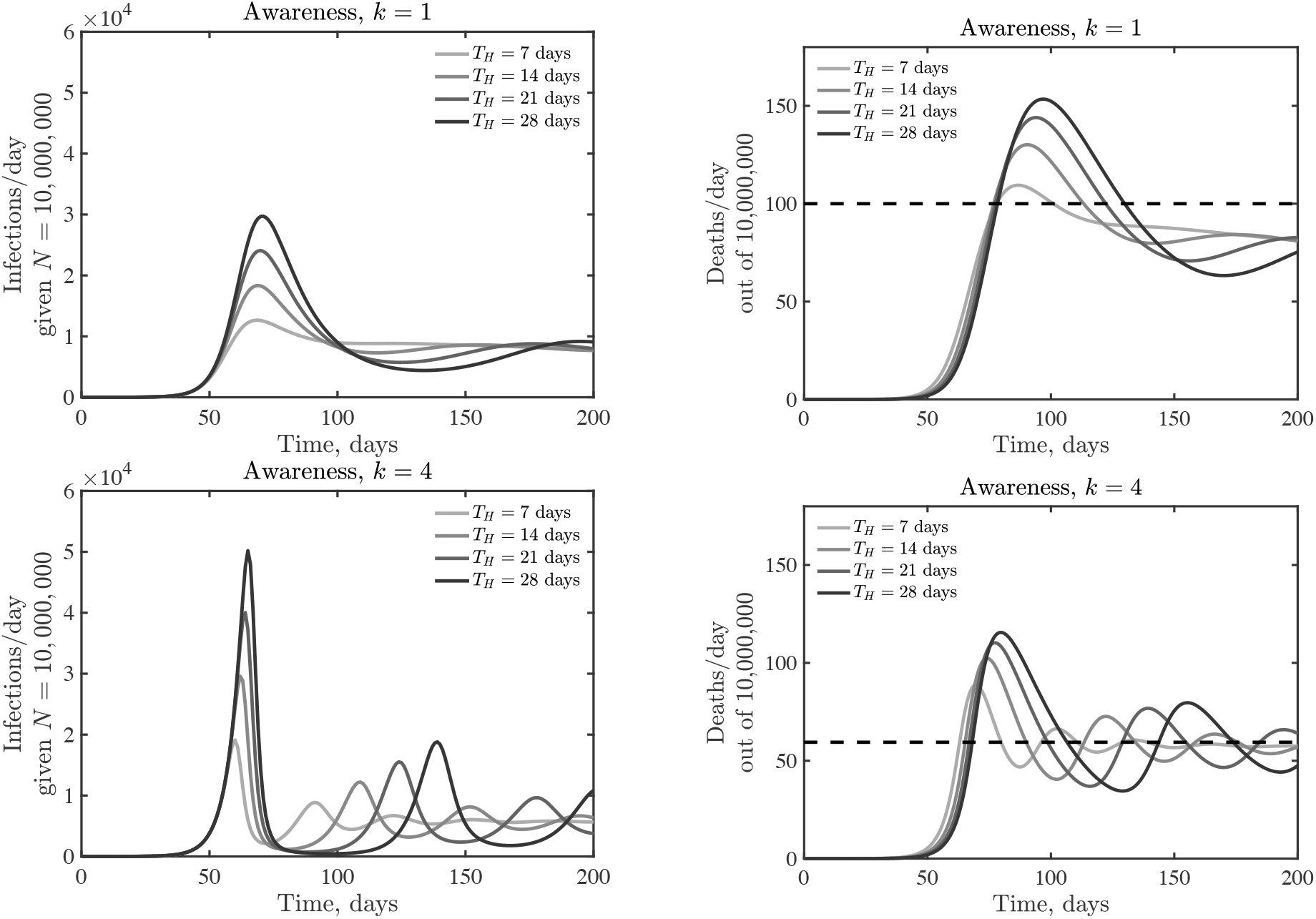
Emergence of oscillatory dynamics in a death-driven awareness model of social distancing given lags between infection and fatality. Awareness is *k* = 1 (top) and *k* = 4 (bottom), all other parameters as in Figure 3. The dashed lines for fatalities expected quasi-stationary value *δ*^(*q*)^.

**FIG. S4:**
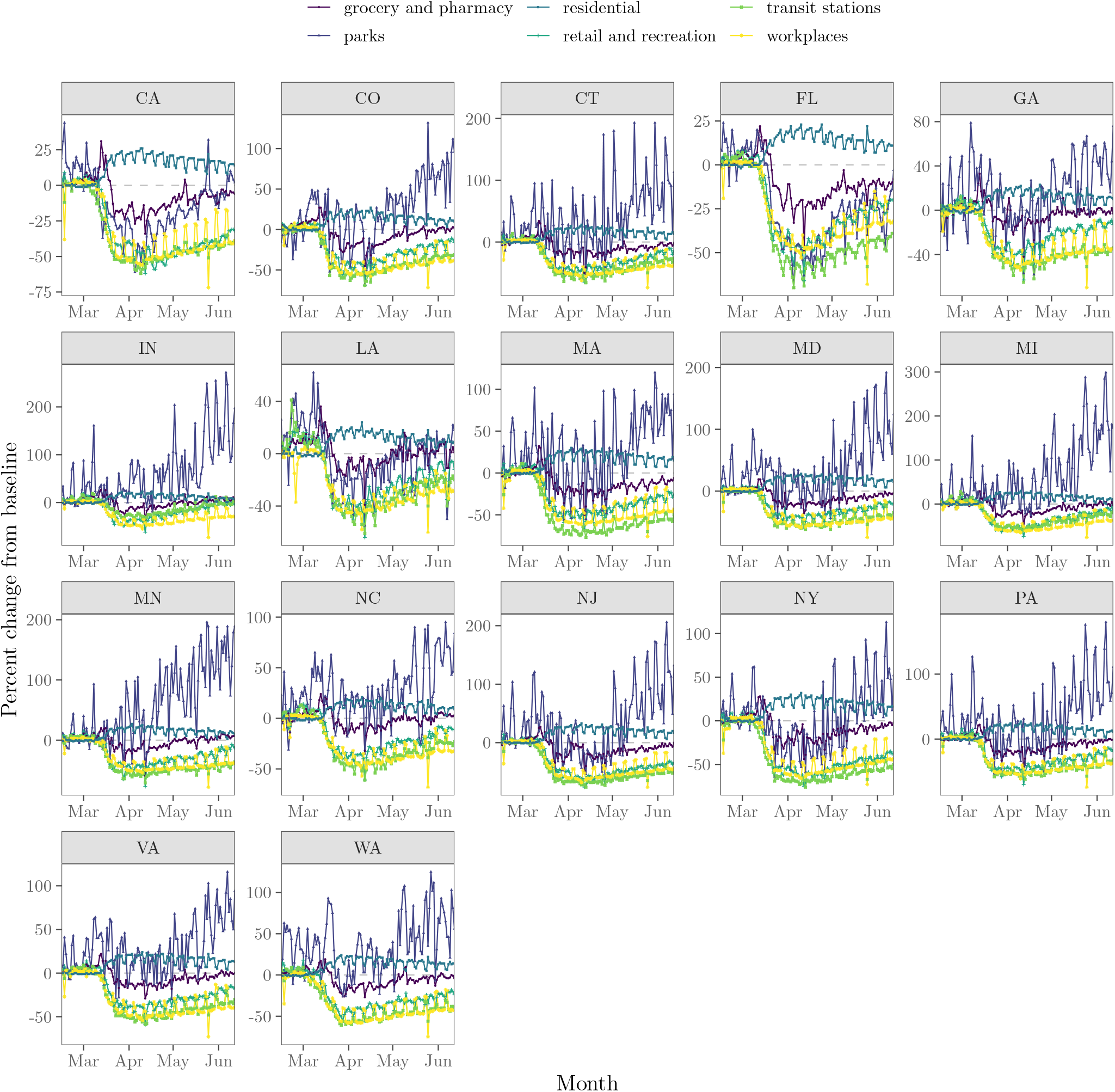
Percent mobility change from baseline across six categories in 17 states.

## Notes

### Competing Interest Statement

The authors have declared no competing interest.

### Summary of Updates

Updated data analysis of asymmetry in fatality data; new model addition to include fatigue; model-data comparison.

